# Diagnostic accuracy of the BD MAX^TM^ MDR-TB assay on sputum and tongue swabs for *Mycobacterium tuberculosis* complex detection in adults under investigation for TB in South Africa

**DOI:** 10.1101/2025.06.10.25329360

**Authors:** Anura David, Lyndel Singh, Manuel Pedro da Silva, Keneilwe Peloakgosi-Shikwambani, Zanele Nsingwane, Violet Molepo, Wendy Stevens, Lesley Erica Scott

## Abstract

**Background:** Despite advances in molecular diagnostics, only 48% of newly diagnosed tuberculosis (TB) cases were confirmed using nucleic acid amplification tests (NAATs) in 2023. The BD MAX^TM^ MDR-TB (MAX MDR-TB) assay, a moderate complexity NAAT, detects *Mycobacterium tuberculosis* complex (MTBC) and resistance to rifampicin (RIF) and isoniazid (INH), but data on clinical performance is limited. This study assessed assay performance on raw sputum, NALC/NaOH decontaminated sputum, and tongue swab (TS) specimens.

**Methods:** This evaluation assessed the MAX MDR-TB assay for MTBC detection and RIF and INH resistance profiling on sputum, using liquid culture as the reference standard. Additionally, diagnostic accuracy for MTBC detection in TS specimens was evaluated under different transport and processing conditions.

**Results:** Assay sensitivity was similar for sputum pellet (87%) and raw sputum (89%), with one additional case detected using raw sputum. Two false RIF-resistant results were observed. INH resistance was missed in two cases. Although specimen numbers were small, TS demonstrated better diagnostic accuracy when using a diluted (66%) STR buffer. A total of 15/55 (27%) were classified as “MTB Low POS.”

**Conclusion:** These findings suggest that MAX MDR-TB assay performance is comparable between sputum pellet and raw sputum. While TS showed promise, further validation in larger studies is warranted. The high rate of “MTB Low POS” results across specimen types underscores the importance of assay optimisation to reduce the burden of repeat testing and improve diagnostic reliability. Future research should enhance sensitivity and integration into diagnostic algorithms to improve patient outcomes.

## Introduction

In 2023, tuberculosis (TB) reclaimed its position as the leading infectious cause of death worldwide, causing an estimated 1.25 million deaths (1). Each year, more than 10 million people are newly infected with diagnosis remaining the weakest link in the TB care cascade (2). Nucleic acid amplification tests (NAATs), such as the GeneXpert MTB/RIF (Xpert) and GeneXpert MTB/RIF Ultra (Ultra) (Cepheid, Sunnyvale, CA), have significantly improved TB diagnosis with implementation of NAATs showing incremental increases over the years. Despite this progress, in 2023, only 48% of newly diagnosed TB cases were bacteriologically confirmed using a NAAT (1). This underscores the need for broader adoption of molecular diagnostics, many of which are able to detect drug resistance in addition to TB, enabling timely patient management (3).

Apart from Molecular WHO recommended rapid diagnostics (mWRD) and culture, sequencing technologies can be used for drug resistance detection, but their high costs and infrastructure demands currently limit their widespread use. While efforts to reduce these costs and make sequencing more accessible are in progress, the rapidly emerging diagnostic pipeline, with options across the laboratory spectrum (from point of care to highly centralized testing) (4) provide opportunities for more countries to adopt molecular testing.

The WHO-recommended moderate complexity assays (3) enable upfront resistance testing for isoniazid (INH) alongside rifampicin (RIF), facilitating informed treatment decisions at the time of diagnosis. These assays were initially evaluated by our group using spiked sputum (5) and these results in conjunction with the WHO-recommendation led to diversification of the diagnostic landscape in South Africa’s (SA’s) TB program (6). However, there is limited clinical data available for these moderate complexity assays.

One of these moderate complexity assays, the BD MAX^TM^ Multi Drug Resistant Tuberculosis (MAX MDR-TB) assay (Becton, Dickinson and Company, Sparks, MD, USA) was incorporated in the South African TB testing algorithm in 2023 (6). This assay is an automated diagnostic test conducted on the BD MAX^TM^ instrument (Becton, Dickinson and Company, Sparks, MD, USA) for detecting *Mycobacterium tuberculosis* complex (MTBC) DNA in raw sputum or sputum pellets (sputum that has been homogenized and decontaminated with NALC-NaOH (N-acetyl-L-cysteine sodium hydroxide)). In specimens where MTBC DNA is identified, the assay detects mutations in the *rpoB* gene associated with RIF resistance, as well as mutations in the *katG* gene and the *inhA* promoter region, both linked to INH resistance. Clinical performance of the MAX MDR-TB assay on respiratory specimens, reported in literature, demonstrates sensitivity ranging from 80% to 91% and specificity from 97% to 100% (7–9), though some studies were limited by small sample sizes. To the authors’ knowledge, there is only one study, published in 2020, which reported an assay sensitivity of 93% in a South African population, with a 32% HIV-positivity rate (10).

In addition to new innovative technologies, efforts to increase access to TB testing has led to the investigation of additional specimen types with oral swabs showing improved sensitivity in recent years (11). Downstream testing of oral swabs is typically performed by “in-house” polymerase chain reaction (PCR) or WHO-recommended low complexity assays (Xpert, Ultra and Truenat MTB (Molbio Diagnostics, Goa, India). Performance data on oral swabs, specifically tongue swabs (TS) on other WHO-recommended NAATs are not available.

In this clinical performance evaluation, we aimed to assess the diagnostic accuracy of the MAX MDR-TB assay on sputum for MTBC, RIF and INH detection compared to a liquid culture reference in a population with a high HIV-positivity rate. Additionally, we assessed the diagnostic accuracy of the MAX MDR-TB assay for MTBC detection in TS specimens (off-label use).

## Methods

### Study design

This cross-sectional, prospective, clinical diagnostic accuracy study evaluated the performance of the MAX MDR-TB assay for MTBC, RIF and INH detection on sputum. Results were compared to a Mycobacterial Growth Indicator Tube (MGIT) (Becton, Dickinson and Company, Sparks, MD, USA) liquid culture reference. In addition, concordance of the MAX MDR-TB assay on TS was assessed using Ultra, liquid culture and MAX MDR-TB sputum results as comparators.

### Ethics statement

Ethics approval for this study was obtained from the University of the Witwatersrand Human Research Ethics Committee (M1911150). The trial was registered with the South African National Clinical Trials Registry (DOH-27-052021-5442). All participants provided written informed consent.

### Participant recruitment and study procedures

Symptomatic adults (≥18 years), being assessed for TB, were enrolled from 20 October 2021 to 11 July 2023 at the Hillbrow Community Health Centre (HCHC), Johannesburg, SA. The WHO-recommended four-symptom screen (W4SS)—cough, fever, weight loss, and night sweats—was administered to participants. Anthropometric characteristics including HIV status, TB history, and demographic information were also collected. Participants’ body mass index (BMI) was also calculated to determine malnutrition, with a BMI <18.5 indicating undernourishment. To be eligible, participants had to agree to a follow-up visit, provide the required specimens, and have no TB treatment in the 6 months prior to enrolment. Specimen collection occurred over two visits (Figure 1); two sputum samples were collected at least 30 minutes apart, with participants refraining from oral intake for at least 30 minutes prior. Two TS were collected using the procedure described by Andama et al. (12). Swab collection from each participant was performed by research nurses by swabbing the dorsum of the tongue with a Copan FLOQSwab (Copan, Brescia, Italy) for 30 seconds, as far back as possible without initiating a gag reflex. One TS was collected before sputum collection and a second swab after sputum collection. After collection, both TS were randomly assigned to tubes containing either 1 mL Tris-EDTA (TE) buffer (10 mM Tris-HCl with 1 mM EDTA Na₂, pH 8.0) (Merck, Johannesburg, SA) or no buffer for transport and storage. Sputum were randomized for routine or research testing.

**Figure 1:**
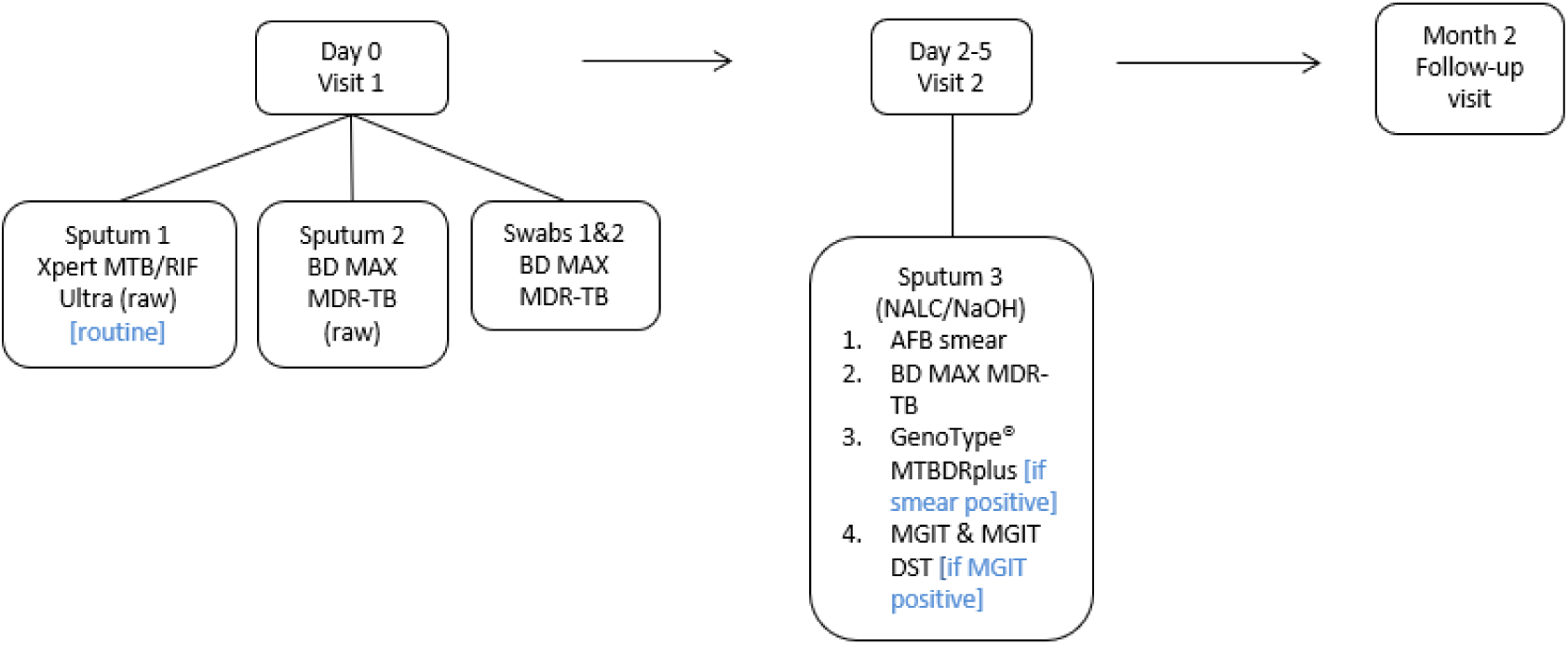
Description of study outline indicating clinic visits and specimen laboratory pathways AFB, acid-fast bacilli; MGIT, Mycobacterial Growth Indicator tube; DST, drug-susceptibility testing; NALC/NaOH,N-acetyl-L-cysteine–sodium citrate–sodium hydroxide; BD, Becton Dickinson

Specimens collected for routine and research testing were transported to the Wits Diagnostic Innovation Hub (WitsDIH) research laboratory in Braamfontein (Johannesburg).

### Routine testing

Only smear-positive sputum (analysed using auramine-O staining) were tested on the Genotype® MTBDR*plus* line probe assay (LPA) (Hain Lifescience/Bruker, Nehren, Germany). All acid-fast bacilli (AFB)-positive cultures were also tested on the LPA to confirm growth of MTBC. Routine testing and result return was performed by laboratory staff as per the National Tuberculosis TB Management Guidelines (13). Staff were blinded to results from the MAX MDR-TB assay.

### MAX MDR-TB assay testing

Raw sputum and sputum pellets were inactivated using a 2:1 ratio of Sample Treatment Reagent (STR) (Becton, Dickinson and Company, Sparks, MD, USA) to specimen before testing on MAX MDR-TB, as per manufacturer instructions. Specimens were batch tested, weekly. For MTBC detection, results are categorized as MTB Detected, MTB Not Detected, MTB Low POS (MTBC DNA detected but resistance metrics not measurable), Indeterminate (due to BD MAX^TM^ system failure), Incomplete (incomplete run), or Unresolved (no MTBC DNA detected, and no internal control detected, indicative of an inhibitory sample or reagent failure). For resistance testing, the assay reports resistance detected (RIF or INH resistance mutations were detected), not detected (RIF or INH resistance mutations were not detected), or unreportable (UNR) (MTBC DNA detected but INH or RIF resistance metrics not measurable). Research staff performed testing and were blinded to routine TB results. Swabs were initially tested using the same processing protocol as that used for sputum (n=271). However, parallel testing on contrived swabs showed improved sensitivity using a diluted (66%) STR buffer, hence 66% STR buffer was used to process the remaining swabs (n=56) (Table 1). Phosphate buffer was used to dilute the STR. Unsuccessful tests were not repeated, as this would compromise the efficiency of rapid diagnostics and incur additional costs.

**Table 1:** Tongue swab processing protocol on the MAX MDR-TB assay

| STR concentration | Swab transport | Volume of STR added to TS |
| --- | --- | --- |
| Neat (n=271) | Dry | 3 mL |
|  | With TE buffer | 2 mL |
| 66% (n=56) | Dry | 3 mL |
|  | With TE buffer | 2 mL |

STR, sample treatment reagent; TS, tongue swab

### Outcomes and statistical analysis

For performance evaluation, raw sputum and sputum pellet results from MAX MDR-TB were compared with liquid culture for MTBC detection and to phenotypic drug susceptibility testing (pDST) (performed using the MGIT960 SIRE kit (Becton, Dickinson and Company, Sparks, MD, USA)) for RIF and INH resistance detection. Data analysis included calculation of sensitivity, specificity, positive predictive value (PPV) and negative predictive value (NPV) for MTBC detection and concordance for resistance detection, with 95% CIs calculated using the Wilson score method (first including all participants and then excluding participants with an Ultra “trace” result). To assess the diagnostic accuracy of the MAX MDR-TB assay on TS, concordance with Ultra, liquid culture and MAX MDR-TB sputum is reported. The target sample size was 400 participants which was derived using the formula described below with population proportion estimated at 50% and 95% confidence interval set at 5%. Sample size (n) = (Z)^2*p(1-p) / x^2, Where Z = 1.96, & where p = population proportion & where x = confidence interval as a proportion. In determining the sample size, allowance was made for participants who do not meet the inclusion criteria. To determine the performance of the MAX MDR-TB assay, only specimens that generated valid results across all tests (MAX MDR-TB, Ultra and MGIT) were included in the statistical analysis.

### Follow up visit

Approximately eight weeks after enrolment, participants were contacted telephonically for follow up. The research nurse evaluated their health status through a series of questions. If the participant reported that they still felt unwell, they were requested to return to HCHC for a follow-up sputum to be collected. Routine testing (smear, MGIT and any other confirmatory testing) was performed.

## Results

### Study population characteristics

Of all participants screened for enrolment over a 21-month period, 133/554 (24%) were found to be ineligible mainly due to a non-productive cough (Figure 2). An additional 86 participants were either lost to follow up or produced unsuccessful test results (either on Ultra, MAX MDR-TB sputum or MGIT). Statistical analysis was therefore performed on 335 participants.

**Figure 2:**
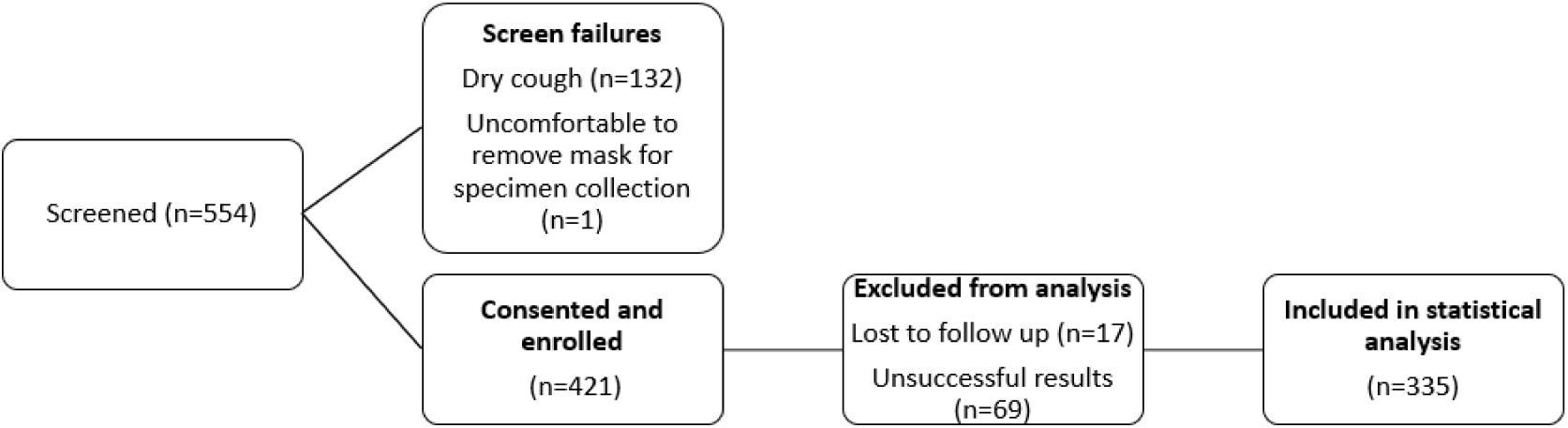
Data description for statistical analysis

The average age of participants was 39 years and 213/335 (64%) were male (Table 2). A total of 186/332 (56%) were HIV-positive with three participants reporting unknown HIV status while 27/62 (44%) participants diagnosed with active TB were malnourished. Previous TB was reported by 42/335 (13%) participants. Cough was the most reported symptom, experienced by 334/335 (99%) of participants with fever being the least reported on 225/335 (67%). Per bacteriological classification using culture, 62/335 (19%) were diagnosed with active TB disease. All participants who were smear-positive (all scanty positive) but culture-negative showed “MTB not detected” results on Ultra, with only one reporting previous TB. The culture contamination rate was 11% (44/404); of these, Ultra and MAX MDR-TB detected one MTBC-positive specimen, and the participant reported an improvement in symptoms after receiving treatment.

**Table 2:**
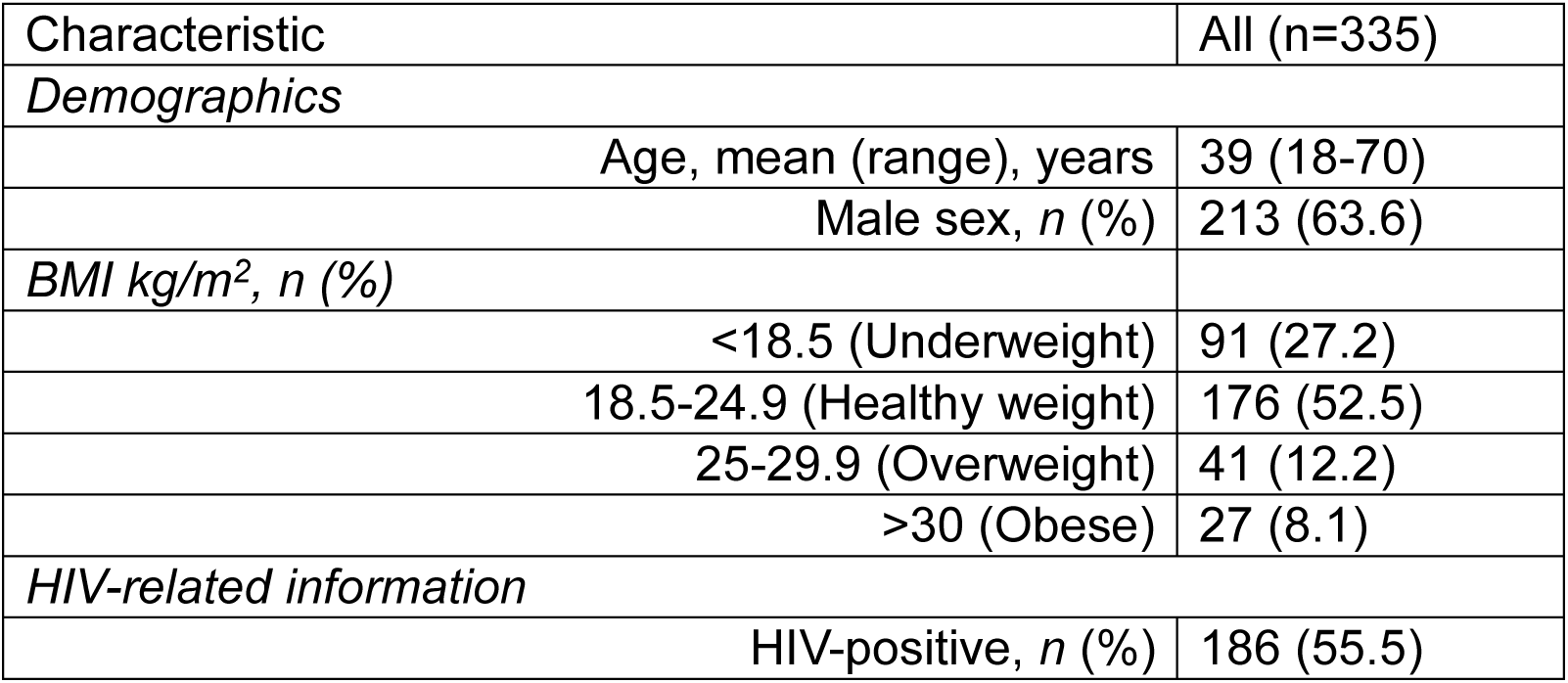
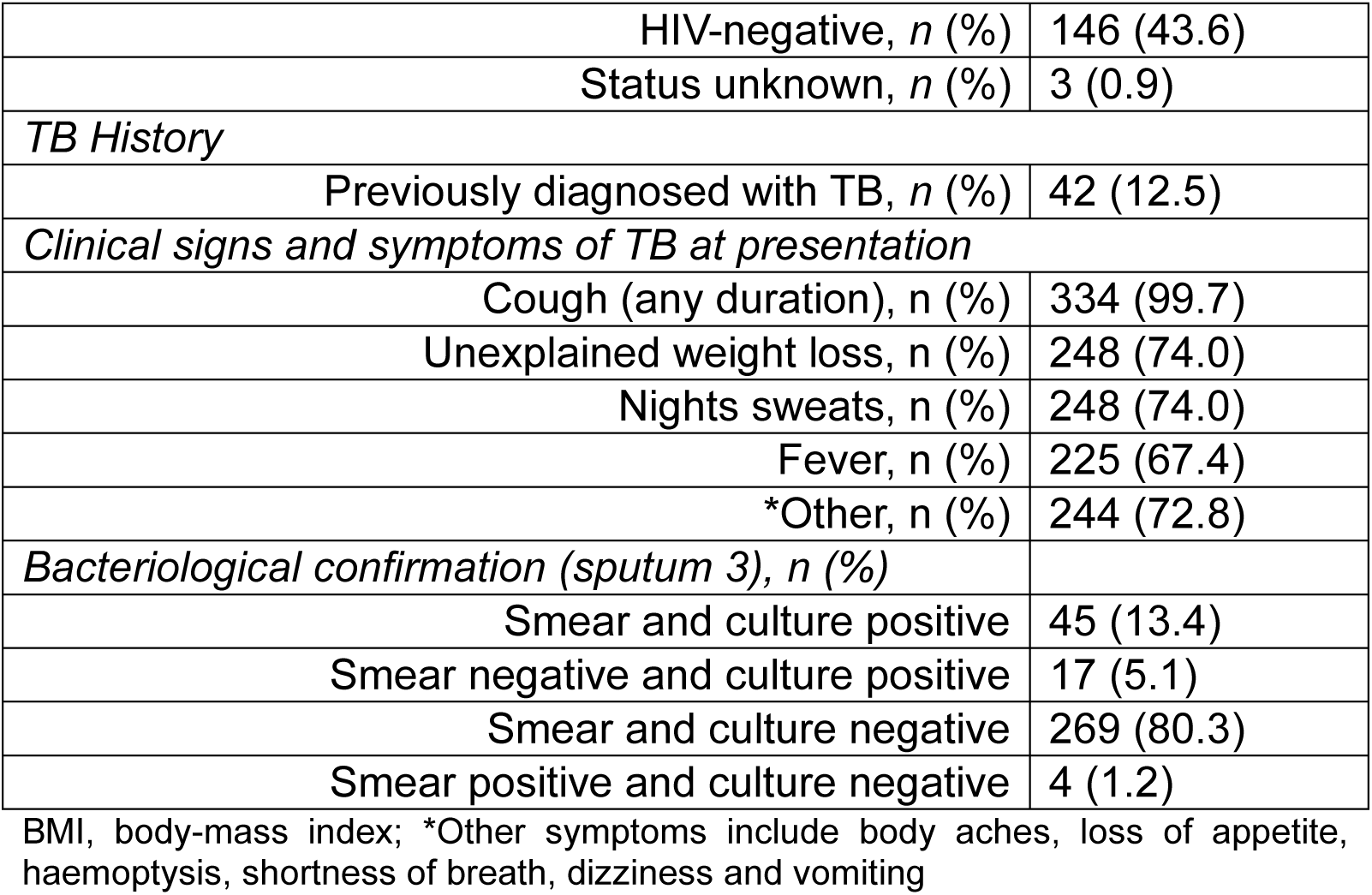
Cohort characteristics for participants used in the statistical analysis

| Characteristic | All (n=335) |
| --- | --- |
| **Demographics** |  |
| Age, mean (range), years | 39 (18-70) |
| Male sex, *n* (%) | 213 (63.6) |
| **BMI kg/m<sup>2</sup>, *n* (%)** |  |
| <18.5 (Underweight) | 91 (27.2) |
| 18.5-24.9 (Healthy weight) | 176 (52.5) |
| 25-29.9 (Overweight) | 41 (12.2) |
| >30 (Obese) | 27 (8.1) |
| **HIV-related information** |  |
| HIV-positive, *n* (%) | 186 (55.5) |
| HIV-negative, *n* (%) | 146 (43.6) |
| Status unknown, *n* (%) | 3 (0.9) |
| *TB History* |  |
| Previously diagnosed with TB, *n* (%) | 42 (12.5) |
| *Clinical signs and symptoms of TB at presentation* |  |
| Cough (any duration), *n* (%) | 334 (99.7) |
| Unexplained weight loss, *n* (%) | 248 (74.0) |
| Nights sweats, *n* (%) | 248 (74.0) |
| Fever, *n* (%) | 225 (67.4) |
| *Other, *n* (%) | 244 (72.8) |
| *Bacteriological confirmation (sputum 3), *n* (%)* |  |
| Smear and culture positive | 45 (13.4) |
| Smear negative and culture positive | 17 (5.1) |
| Smear and culture negative | 269 (80.3) |
| Smear positive and culture negative | 4 (1.2) |

BMI, body-mass index; \*Other symptoms include body aches, loss of appetite, haemoptysis, shortness of breath, dizziness and vomiting

### Diagnostic evaluation of the MAX MDR-TB assay on sputum for MTBC detection

The performance of MAX MDR-TB on raw sputum and sputum pellets was compared with liquid culture and, additionally, stratified by HIV and smear status, as outlined in Table 3 for 335 specimen results. Out of the 55 positive MTBC results reported by the MAX MDR-TB assay on raw sputum, 15 (27%) were classified as “MTB Low POS.” For the five raw sputum samples in which MTBC was detected using the MAX MDR-TB assay but not by culture, MTBC was detected by other NAATs in two participants, one by LPA and the other by Ultra. None of the five participants reported previous TB. For the four sputum pellets where MTBC was detected on the MAX MDR-TB assay but not on culture, MTBC was detected on Ultra for one participant. Of these participants, 3/4 (75%) reported previous TB. One of these participants with a positive MAX MDR-TB but negative culture result could not be contacted at the 8-week follow-up visit while the other eight reported an improvement in symptoms, without TB treatment. Unsuccessful results (either indeterminate or UNR) were reported on the raw sputum and/or sputum pellet on 25/403 (6%) participants. The MAX MDR-TB assay yielded a “UNR” result on both specimen types (raw sputum and pellet) for 1/25 (4%) participant.

**Table 3:** The performance of smear microscopy, Ultra and MAX MDR-TB assays on sputum compared to the liquid culture for MTBC detection

| Variable | Smear microscopy | Ultra (raw sputum) | MAX MDR-TB (raw sputum) | MAX MDR-TB (sputum pellet) |
| --- | --- | --- | --- | --- |
| *All (including trace) (n=335)* |  |  |  |  |
| Sensitivity, % (95% CI) | 72.6 (59.8-85.4) | 95.2 (86.5-99.9) | 88.7 (78.1-95.3) | 87.1 (76.1-94.3) |
| Specificity, % (95% CI) | 98.5 (96.3-100) | 97.8 (95.3-99.2) | 98.2 (98.6-100) | 98.5 (96.3-99.6) |
| PPV, % (95% CI) | 88.2 (76.1-100) | 90.8 (81.0-96.5) | 91.7 (81.6-97.2) | 93.1 (83.3-98.1) |
| NPV, % (95% CI) | 93.0 (89.2-96.8) | 98.9 (96.8-99.8) | 97.5 (94.8-99.0) | 97.1 (94.4-98.7) |
| *All (excluding trace) (n=329)* |  |  |  |  |
| Sensitivity, % (95% CI) | 73.3 (60.3-86.3) | 95.0 (86.1-99.0) | 91.7 (81.6-97.2) | 88.3 (77.4-95.2) |
| Specificity, % (95% CI) | 98.5 (96.2-100) | 99.3 (97.3-99.9) | 98.1 (95.7-99.4) | 98.9 (96.8-99.8) |
| PPV, % (95% CI) | 91.8 (80.4-100) | 96.6 (88.3-99.6) | 91.7 (81.6-97.2) | 94.6 (85.1-98.9) |
| NPV, % (95% CI) | 94.1 (90.7-97.5) | 98.9 (96.8-99.8) | 98.1 (95.7-99.4) | 97.4 (94.8-99.0) |
| *Specimens from HIV-positive individuals (n=186)* |  |  |  |  |
| Sensitivity, % (95% CI) | 62.5 (43.7-81.3) | 93.8 (79.2-99.2) | 78.1 (60.0-90.7) | 78.1 (60.0-90.7) |
| Specificity, % (95% CI) | 98.7 (95.4-100) | 97.4 (93.5-99.3) | 98.7 (95.4-99.8) | 98.1 (94.4-99.6) |
| PPV, % (95% CI) | 91.7 (80.0-100) | 88.2 (72.5-96.7) | 92.6 (75.7-99.7) | 89.3 (71.8-97.7) |
| NPV, % (95% CI) | 94.3 (90.9-97.7) | 98.7 (95.3-99.8) | 95.6 (91.1-98.2) | 95.6 (91.1-98.2) |
| *Specimens from HIV-negative individuals (n=146)* |  |  |  |  |
| Sensitivity, % (95% CI) | 85.7 (67.3-100) | 96.4 (81.7-99.9) | 100 (87.7-100) | 96.4 (81.7-99.9) |
| Specificity, % (95% CI) | 98.3 (94.0-100) | 98.3 (94.0-99.8) | 97.5 (92.7-99.5) | 99.2 (95.4-100) |
| PPV, % (95% CI) | 92.3 (74.9-100) | 93.1 (77.2-99.2) | 90.3 (74.2-98.0) | 96.4 (81.7-99.9) |
| NPV, % (95% CI) | 96.7 (91.7-100) | 99.1 (95.3-100) | 100 (96.8-100) | 99.2 (95.4-100) |
| *Smear microscopy negative specimens (n=286)* |  |  |  |  |
| Sensitivity, % (95% CI) | n/a | 88.2 (63.6-98.5) | 70.6 (44.0-89.7) | 64.7 (38.3-85.8) |
| Specificity, % (95% CI) |  | 97.8 (95.2-99.2) | 98.5 (96.2-99.6) | 98.5 (96.2-99.6) |
| PPV, % (95% CI) |  | 71.4 (47.8-88.7) | 75.0 (47.65-92.7) | 73.3 (44.9-92.2) |
| NPV, % (95% CI) |  | 99.2 (97.3-99.9) | 98.1 (95.7-99.4) | 97.8 (95.2-99.2) |

PPV, positive predictive value; NPV, negative predictive value; CI, Confidence interval; Ultra refers to the Xpert MTB/RIF Ultra assay

### Comparison of the MAX MDR-TB assay with the Ultra assay for MTBC detection

In this study population, when compared with the liquid culture reference, Ultra demonstrated a sensitivity of 95%, while the MAX MDR-TB assay showed a sensitivity of 89%, with Ultra detecting TB in four additional participants. Specificities between the two assays were similar. On raw sputum, there were 4/404 (<1%) unsuccessful results on Ultra compared to 19/403 (5%) unsuccessful results on MAX MDR-TB.

### Detection of drug resistance

#### RIF resistance profiling sputum on the Ultra, liquid culture and MAX MDR-TB assay

Rifampicin results for Ultra, MAX MDR-TB and culture are shown in Figure 3. Unsuccessful RIF results were produced on 6/65 (9%), 6/62 (10%) and 24/60 (40%) sputum for Ultra, culture and MAX MDR-TB, respectively.

**Figure 3:**
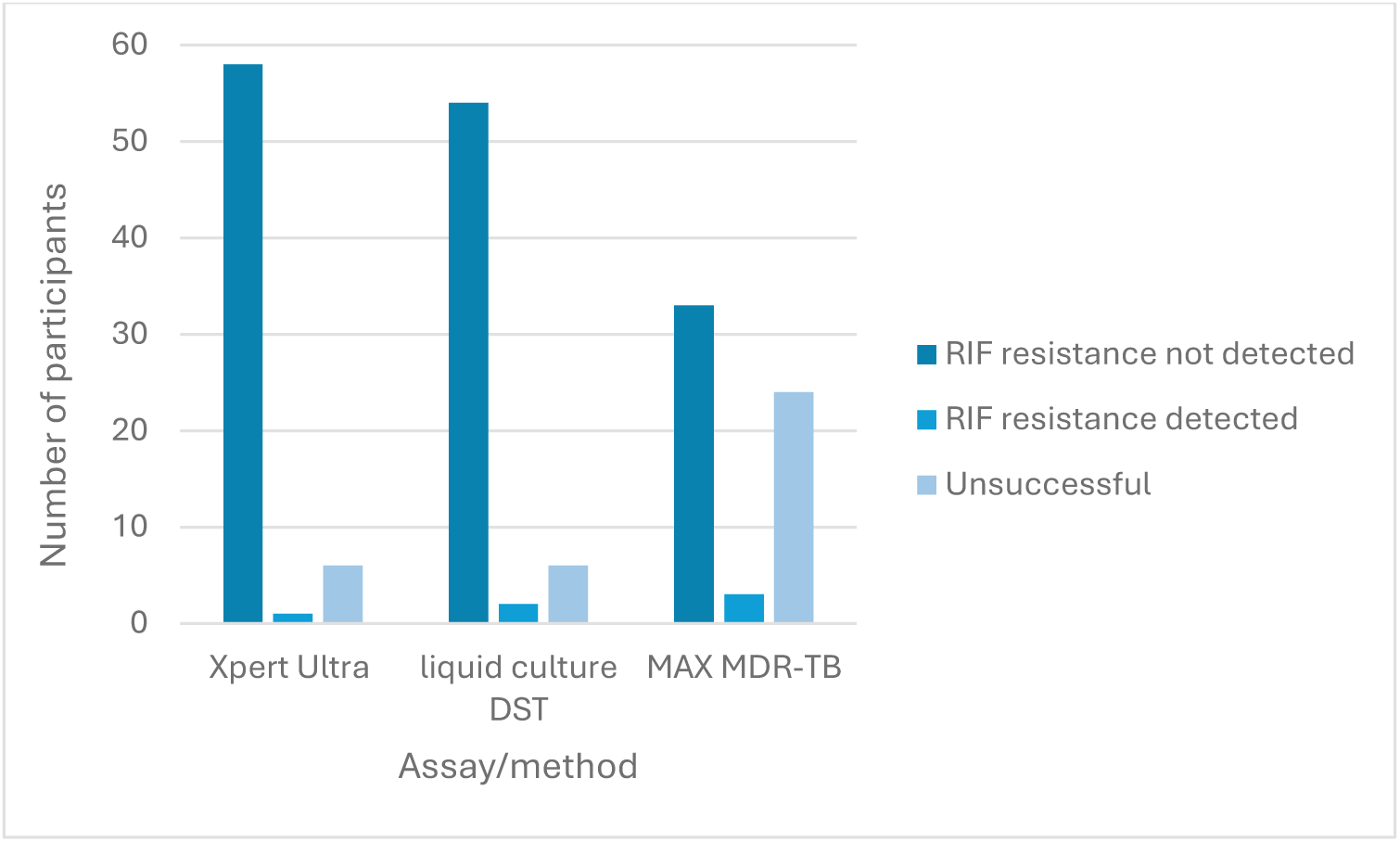
RIF resistance profiles for the Ultra assay, liquid culture and MAX MDR-TB assay RIF, rifampicin; DST, drug susceptibility testing, Ultra refers to the Xpert MTB/RIF Ultra assay, MAX MDR-TB refers to the BD MAX^TM^ MDR-TB assay

On the MAX MDR-TB assay, valid RIF resistance profiles were available for 35/55 (63%) sputum. When compared to pDST for RIF, the MAX MDR-TB assay, on sputum, correctly assigned resistance profiles to 31/33 (94%) participants (Table 4). Two raw sputum specimens which did not show any resistance on pDST, LPA, MAX MDR-TB sputum pellet or on the Ultra assay were identified as resistant by the MAX MDR-TB assay. Sputum from three participants produced valid RIF results, but INH results were reported as “unreportable.”

**Table 4:** MAX MDR-TB rifampicin resistance results, compared to phenotypic pDST

| pDST result | MAX MDR-TB result |  |  |  |
| --- | --- | --- | --- | --- |
|  | RIF-R not detected | RIF-R detected | Indeterminate | Unreportable |
| **RIF sensitive**<br>(n=48) | 30 | 2 | 11 | 5 |
| **RIF resistant (n=1)** | 0 | 1 | 0 | 0 |
| **Unsuccessful**<br>(n=6) | 2 | 0 | 4 | 0 |

RIF, rifampicin; RIF-R, rifampicin resistance; pDST, phenotypic drug susceptibility testing; Unsuccessful pDST results refer to cultures which failed to grow in the control tube or were repeatedly contaminated

#### INH resistance profiling for sputum on the MAX MDR-TB assay

Valid INH resistance profiles were available for 38/55 (69%) sputum (Table 5). When compared to pDST for INH the MAX MDR-TB assay, on sputum, correctly assigned resistance profiles to 34/36 (94%) participants. INH resistance was not detected by MAX MDR-TB on either raw sputum or sputum pellet for two participants, but resistance was detected by pDST. INH resistance was also not detected on the LPA for these participants. Unsuccessful INH results were reported in 17/55 (31%) tests performed. Sputum from five participants produced valid INH results, but RIF results were reported as “unreportable.”

**Table 5:** MAX MDR-TB isoniazid resistance results, compared to phenotypic DST

| pDST result | MAX MDR-TB result |  |  |  |
| --- | --- | --- | --- | --- |
|  | INH-R not detected | INH-R detected | Indeterminate | Unreportable |
| **INH sensitive**<br>(n=45) | 33 | 0 | 10 | 2 |
| **INH resistant (n=4)** | 2 | 1 | 1 | 0 |
| **Unsuccessful (n=6)** | 2 | 0 | 4 | 0 |

INH, isoniazid; INH-R, isoniazid resistance; pDST, phenotypic drug susceptibility testing; Unsuccessful pDST results refer to cultures which failed to grow in the control tube or were repeatedly contaminated

#### Diagnostic accuracy of the MAX MDR-TB assay on tongue swabs

A total of 320 TS were tested on the MAX MDR-TB assay. For those TS processed using neat STR and found to be MTBC-positive, 12/19 (63%) and 26/32 (81%) transported “dry” and in TE buffer, respectively, were classified as “MTB Low POS.” When neat STR was used for TS processing, improved detection of MTBC was observed on TS transported in TE buffer compared to those transported “dry,” regardless of the comparator test (Table 6). When processed with neat STR, TS transported in TE buffer showed MTBC detection in specimens down to a semi-quantitative “low” Ultra whereas TS without buffer only detected MTBC in samples with a semi-quantitative result of “medium” or higher. Although neat STR processing demonstrated reduced specificity when TS were transported in TE buffer, they showed a reduced error rate (3% [11/320] compared to 7% [21/320]) for TS that were transported “dry.” Both sensitivity and specificity seemed improved when using 66% STR but the sample size was limited on MTBC-positive TS. No MTBC detection was seen on TS from participants who produced a trace sputum result on Ultra (n=6). Of these, 2/6 (33%) were confirmed to be bacteriologically positive by culture.

**Table 6:** Performance of the MAX MDR-TB assay on tongue swabs compared to Ultra, liquid culture and MAX MDR-TB sputum results

| Comparator test result | MAX MDR-TB TS result, n/N (%) |  |  |  |
| --- | --- | --- | --- | --- |
|  | Dry | With TE buffer | Dry | With TE buffer |
|  | Neat STR | Neat STR | 66% STR | 66% STR |
| **Ultra positive** | 12/50 (24.0) | 20/52 (38.5) | 6/9 (66.7) | 6/9 (66.7) |
| *High* | 10/18 (55.6) | 13/19 (68.4) | 2/4 (50.0) | 2/4 (50.0) |
| *Medium* | 2/6 (33.3) | 3/6 (50.0) | 1/1 (100) | 1/1 (100) |
| *Low* | 0/13 (0) | 4/14 (28.6) | 3/3 (100) | 3/3 (100) |
| *Very low* | 0/7 (0) | 0/7 (0) | 0/1 (0) | 0/1 (0) |
| *Trace* | 0/6 (0) | 0/6 (0) | - | - |
| **Ultra negative** | 201/202 (99.5) | 204/210 (97.1) | 38/38 (100) | 38/38 (100) |
| **Liquid culture positive** | 13/47 (27.7) | 21/50 (42.0) | 6/8 (75.0) | 6/8 (75.0) |
| **Liquid culture negative** | 205/205 (100) | 207/212 (97.6) | 39/39 (100) | 39/39 (100) |
| **MAX MDR-TB sputum positive** | 13/46 (28.3) | 21/48 (43.8) | 6/8 (75.0) | 6/8 (75.0) |
| **MAX MDR-TB sputum negative** | 206/206 (100) | 209/214 (97.7) | 39/39 (100) | 39/39 (100) |

STR, sample treatment reagent; TS, tongue swabs; Ultra refers to the Xpert MTB/RIF Ultra assay

When TS results were stratified by collection time relative to sputum collection, independent of TE buffer use or laboratory processing method, MTBC was detected in 24/320 (8%) specimens collected prior to sputum collection and in 27/320 (8%) specimens collected afterward.

## Discussion

This study evaluated the clinical performance of the MAX MDR-TB assay on raw sputum and sputum pellet for MTBC, RIF and INH detection compared to a liquid culture reference. Additionally, we assessed the diagnostic accuracy of the MAX MDR-TB assay for MTBC detection on TS specimens.

In this study, ∼44% of participants with active TB were undernourished as indicated by their Body Max Index (BMI) which is a TB risk factor TB (1) and is also associated with increased TB incidence and severity (14).

Assay sensitivity was similar between the sputum pellet (87%) and raw sputum (89%) with MTBC being detected on one additional raw sputum. This falls in the upper end of sensitivity reported in literature (7, 10). On raw sputum, Ultra, demonstrated better performance (95%) than the MAX MDR-TB assay, identifying TB in four additional participants. Similar findings were published by Mokaddas *et al* (15) where Ultra diagnosed eight additional participants compared to the MAX MDR-TB assay. Raw sputum from 27% of participants yielded an “MTB low POS” result on the MAX MDR-TB assay which the manufacturer recommends repeating. This translates to an additional cost for the laboratory. For patient management, this means a delayed result and if the repeat test yields the same result, then this individual needs to return to the healthcare facility to provide a second sputum for resistance testing. Similarly, a “trace” result on the Ultra assay, observed in six participants in this study population, also requires follow-up testing. These results suggest that nine additional participants may have missed appropriate treatment initiation if the MAX MDR-TB assay was used as the initial TB diagnostic. The MAX MDR-TB assay demonstrated better sensitivity, on raw sputum, in the HIV-negative cohort (100%) compared to the HIV-positive cohort (78%). This is not surprising since HIV/TB co-infection is linked to lower bacillary loads, making TB diagnosis more challenging (16).

Of the five potential false-positive results identified by the MAX MDR-TB assay, all participants who could be contacted at the 8-week follow-up reported symptomatic improvement in the absence of any treatment intervention.

The MAX MDR-TB assay has the advantage of producing a valid result for one drug while the other may be reported as “unreportable.” For drug-resistance profiling, two false RIF-resistant results were reported by MAX MDR-TB on raw sputum. Some factors contributing to false-positive results can include the detection of non-pathogenic mutations in the *rpoB* gene (17), technical issues within the assay, or mixed infections. Both the MAX MDR-TB assay and the LPA missed INH-resistance on two participants. It is possible for phenotypic resistance to be observed before the corresponding genetic mutations are detected. This discrepancy can occur due to limitations in the sensitivity of genotypic assays which typically target the most commonly seen mutations or the presence of resistance mechanisms that are not yet genetically characterized (18). The MAX MDR-TB assay was able to produce valid resistance profiles for two participants whose pDST tests were unsuccessful.

For the MAX MDR-TB performance on TS, diagnostic accuracy was chosen over analytical sensitivity and specificity due to the lack of an optimized TS processing method. When using neat STR, TS transportation in TE buffer appeared to improve the sensitivity of the MAX MDR-TB assay with a reduced specificity compared to those that were transported “dry.” For TS specimens processed with 66% STR buffer, the limited number of MTBC-positive cases precludes drawing definitive conclusions regarding the assay’s ability to detect MTBC. Nonetheless, assay specificity on TS appeared to be higher with 66% STR compared to neat STR, regardless of buffer or “dry” transportation. A notable limitation observed with TS tested on the MAX MDR-TB assay is the high frequency of “MTB Low POS” results. Despite this disadvantage, TS paired with a NAAT could increase access to TB testing, especially in populations who are unable to produce sputum. In this study, 132 participants were ineligible for this study due to a non-productive cough. This population needs to be investigated in future TS evaluation studies as they are likely representative of individuals that are being targeted by initiatives such as Targeted Universal TB Testing (TUTT) (19). An analysis of TS performance, stratified by whether the swab was collected before or after sputum collection, indicates that the timing of TS collection does not impact diagnostic performance. Overall, these findings suggest that TS is a viable specimen type for MTBC detection using the MAX MDR-TB assay. Consistent with observations from WHO-recommended low complexity assays (20, 21), designed for sputum, MTBC detection from TS is improved in individuals with higher bacterial loads, a trend also observed with the MAX MDR-TB assay.

Although point estimates for diagnostic sensitivity and specificity, on sputum, differed between MAX MDR-TB and Ultra, the overlapping confidence intervals suggest similar performance between the assays. There were also fewer unsuccessful results (<1%) seen on the Ultra assay compared to the MAX MDR-TB assay (5%). This highlights the importance of considering the uncertainty around diagnostic performance when selecting assays for clinical implementation. Overall, the MAX MDR-TB assay remains a valuable diagnostic tool, particularly in automated laboratory settings where streamlined workflows are critical. However, the risk of a lower TB yield compared to the Ultra assay underscores the need for further optimization and careful evaluation in diverse clinical settings. Future research should focus on enhancing assay sensitivity and assessing its role within integrated TB diagnostic algorithms to improve patient outcomes.

## Data Availability

All data produced in the present study are available upon reasonable request to the authors

## Acknowledgments

We thank the study participants; the Johannesburg Health District and the and the Gauteng Department of Health for their support and collaboration on this study; Becton, Dickinson and Company for providing technical support, kits and BD MAX^TM^ Systems for this study. Becton, Dickinson and Company was not involved in the study design and analysis and interpretation of results.

## Author contributions

Conceptualization, A.D. and L.E.S.; Data curation, A.D.; Formal analysis, A.D.; Funding acquisition, L.E.S. and W.S.; Investigation, A.D., L.S. K.P-S. Z.N. and V.M.; Methodology, A.D. and L.E.S.; Project administration, L.E.S. and W.S.; Supervision, L.E.S.; Validation, A.D.; Visualization, A.D. and L.E.S. ;Writing—original draft, A.D.; Writing—review and editing, A.D., L.E.S., M.P.d.S., L.S., K.P-S., Z.N., V.M. and W.S. All authors have read and agreed to the published version of the manuscript.

## References

1. Global Tuberculosis Report 2024. World Health Organisation; 2024 Nov [date accessed 04 November 2024]. Available from https://www.who.int/teams/global-tuberculosis-programme/tb-reports/global-tuberculosis-report-2024

2. Pai M, Dewan PK, Swaminathan S. Transforming tuberculosis diagnosis. Nat Microbiol. 2023;8(5):756–9.

3. Module 3: Diagnostics, Rapid Diagnostic for Tuberculosis Detection, WHO consolidated guidelines on Tuberculosis, World Health Organization; 2021 July [date accessed 07 July 2022]. Available from: https://www.who.int/publications/i/item/9789240029415.

4. Pipeline Report 2023. Treatment Action Group. 2023. [date accessed: 15 February 2024]. Available from: https://www.treatmentactiongroup.org/wp-content/uploads/2023/11/2023_pipeline_TB_diagnostics_final.pdf

5. de Vos M, Scott L, David A, Trollip A, Hoffmann H, Georghiou SB, Carmona S, Ruhwald M, Stevens W, Denkinger C, Schumacher S. Comparative analytical evaluation of four centralized platforms for the detection of M. tuberculosis complex and resistance to rifampicin and isoniazid. J Clin Microbiol. 2021;59(3):e02168–20.

6. Yes, we can end TB. 2023. NICD. [date accessed 27 January 2025]. Available from https://www.nicd.ac.za/wp-content/uploads/2023/03/World-TB-Day-2023.pdf. [press release].

7. Ko SJ, Yoon KH, Lee SH. Performance of the BD MAX MDR-TB assay in a clinical setting and its impact on the clinical course of patients with pulmonary tuberculosis: a retrospective before-after study. J Yeungnam Med Sci 2024;41(2):113–9.

8. Hofmann-Thiel S, Plesnik S, Mihalic M, Heiß-Neumann M, Avsar K, Beutler M, Hoffmann H. Clinical Evaluation of BD MAX MDR-TB Assay for Direct Detection of Mycobacterium tuberculosis Complex and Resistance Markers. J Mol Diagn. 2020;22(10):1280–6.

9. Sağıroğlu P, Atalay MA. Evaluation of the performance of the BD MAX MDR-TB test in the diagnosis of Mycobacterium tuberculosis complex in extrapulmonary and pulmonary samples. Expert Rev Mol Diagn. 2021;21(12):1361–7.

10. Shah M, Paradis S, Betz J, Beylis N, Bharadwa jR, Caceres T, Gotuzzo E, Joloba M, Mave V, Nakiyingi L, Nicol MP, Pradhan N, King B, Armstrong D, Knecht D, Maus CE, Cooper CK, Dorman SE, Manabe YC. Multicenter Study of the Accuracy of the BD MAX™ MDR-TB Assay for Detection of Mycobacterium tuberculosis Complex and Mutations Associated with Resistance to Rifampin and Isoniazid. Clin Infect Dis. 2019;71(5):1161–7.

11. Church EC, Steingart KR, Cangelosi GA, Ruhwald M, Kohli M, Shapiro AE. Oral swabs with a rapid molecular diagnostic test for pulmonary tuberculosis in adults and children: a systematic review. Lancet Glob Health. 2024;12:e45–54.

12. Andama A, Steadman AE, Ahls C, Cangelosi GA, David A, de Vos M, Heichman K, Kato-Maeda M, Penn-Nicholson A, Olson A, Scott L, Turnbull L, Wood R, Weigel K, Cattamanchi A. Consensus standard operating procedure for collection of tongue swabs for TB diagnostics 2024 [Available from: https://www.protocols.io/view/consensus-standard-operating-procedure-for-collect-kxygxyw54l8j/v1.

13. South African National Department of Health. National guideline on the treatment of tuberculosis infection. [date accessed: 21 August 2024]. [press release]. 2023.

14. Kumar NP, Nancy AP, Moideen K, Menon PA, Banurekha VB, Nair D, Nott S, Babu S. Low body mass index is associated with diminished cytokines and chemokines in both active and latent tuberculosis. Nutr Immunol. 2023;10.

15. Mokaddas EM, Ahmad S, Eldeen HS. GeneXpert MTB/RIF Is Superior to BBD Max MDR-TB for Diagnosis of Tuberculosis (TB) in a Country with Low Incidence of Multidrug-Resistant TB (MDR-TB). J Clin Microbiol. 2019;57(10).

16. Vittor AY, Garland JM, Schlossberg D. Improving the Diagnosis of Tuberculosis: From QuantiFERON to New Techniques to Diagnose Tuberculosis Infections. Curr HIV/AIDS Rep. 2011;8:153–63.

17. Shea J, Halse TA, Kohlerschmidt D, Lapierre P, Modestil HA, Kearns CH, Dworkin FF, Rakeman JL, Escuyer V, Musser KA. Low-Level Rifampin Resistance and rpoB Mutations in Mycobacterium tuberculosis: an Analysis of Whole-Genome Sequencing and Drug Susceptibility Test Data in New York. J Clin Microbiol. 2021;59(4).

18. Cohen KA, Manson AL, Desjardins CA, Abeel T, Earl AM. Deciphering drug resistance in Mycobacterium tuberculosis using whole-genome sequencing: progress, promise, and challenges. Genome Med. 2019;11(45).

19. Martinson NA, Nonyane BAS, Genade LP, Berhanu RH, Naidoo P, Brey Z, Kinghorn A, Nyathi S, Young K, Hausler H, Connell L, Lutchminarain K, Swe Swe-Han K, Vreede H, Said M, von Knorring N, Moulton LH, Lebina L, team TT. Evaluating systematic targeted universal testing for tuberculosis in primary care clinics of South Africa: A cluster-randomized trial (The TUTT Trial). PLoS Med. 2023;20(5).

20. Andama A, Whitman G, Crowder R, Reza T, Jaganath D, Mulondo J, Nalugwa T, Semitala F, Worodria W, Cook C, Wood R, Weigel K, Olson A, Shaw J, Kato-Maeda M, Denkinger C, Nahid P, Cangelosi G, Cattamanchi A. Accuracy of Tongue Swab Testing Using Xpert MTB-RIF Ultra for Tuberculosis Diagnosis. J Clin Microbiol. 2022;60(7).

21. Wood RC, Luabeya AK, Dragovich RB, Olson AM, Lochner KA, Weigel KM, Codsi R, Mulenga H, de Vos M, Kohli M, Penn-Nicholson A, Hatherill M, Cangelosi GA. Tongue swab testing on two automated tuberculosis diagnostic platforms, Cepheid Xpert® MTB/RIF Ultra and Molbio Truenat® MTB Ultima. J Clin Microbiol. 2024;10(4).

